# Long COVID and the Increased Risk of Food Insecurity Among Participants in Arizona CoVHORT: A matched cohort study

**DOI:** 10.1101/2025.11.14.25340219

**Authors:** Chidera M Ejike, Laura P Falk, Kristen Pogreba-Brown, Kacey Ernst

## Abstract

**Objectives:** To assess whether individuals with long COVID face a higher risk of food insecurity compared with those without long COVID.

**Methods:** We used data from the Arizona CoVHORT, which is a prospective longitudinal study of SARS-CoV-2 health outcomes initiated in May 2020. Participants with confirmed infections who completed a symptom survey ≥6 months post-infection were eligible (n = 2415). We matched participants with long COVID to participants who did not have long COVID by gender, age group, income bracket, and date of first assessment of food insecurity (±6 months). We estimated the association between long COVID and food insecurity using a conditional logistic regression analysis.

**Results:** Participants with long COVID had significantly greater odds of food insecurity (adjusted odds ratio = 1.47; 95% confidence interval = 1.06 - 2.04).

**Conclusions:** Long COVID significantly increases vulnerability to food insecurity. This highlights the need for integrated health and social interventions for individuals with long COVID.

**WHAT IS ALREADY KNOWN ON THIS TOPIC:** Food Insecurity has been shown to be an important risk factor for long COVID, but we do not know whether long COVID independently increases the risk of food insecurity, nor have studies adopted a cohort design approach to examine the relationship between long COVID and food insecurity. This study is the first to analyze the association between long COVID and food Insecurity using a matched cohort data within the United States.

**WHAT THIS STUDY ADDS:** This study uniquely adds to the current knowledge base by providing evidence that long COVID is independently associated with a significantly increased risk of food insecurity among adults in Arizona, after adjusting for confounders. We found that adults with long COVID in the Arizona CoVHORT had 47% higher odds of experiencing food insecurity compared to matched controls without long COVID. This study provides robust statistical support for a direct association between long COVID and food insecurity and advances the field by evidencing a previously under-explored dimension of the pandemic’s long-term impact, thus, highlighting an urgent need for integrated screening and intervention strategies at the intersection of chronic illness and nutrition insecurity.

**HOW THIS STUDY MIGHT AFFECT RESEARCH, PRACTICE OR POLICY:** Our study provides evidence that long COVID not only affects health of individuals but also social and economic well-being, including food access and stability. Healthcare and public health systems should integrate routine screening for food insecurity in outpatient care, primary care, and long COVID clinics. Early detection and referral to nutritional or financial assistance programs may reduce the combined effects of chronic illness and economic strain. These interventions are critical for groups disproportionately impacted by the pandemic’s longer-term effects, including low-income households, women, and communities of color. Expanding safety net programs and tracking social consequences of infectious diseases are vital for public health preparedness.

## INTRODUCTION

The National Academies of Sciences, Engineering, and Medicine defines long COVID (LC) as an infection-associated chronic condition that occurs after SARS-CoV-2 infection and persists for at least three months as a continuous, relapsing-remitting, or progressive disease affecting one or more organ systems.(1) Long COVID affects an estimated 6.4-14.7% of the U.S. population,(2,3) presenting with diverse symptoms, the onset or worsening of chronic conditions, and limitations in daily activities.(1,4) These health impacts can disrupt employment and income,(5) creating both financial and nonfinancial barriers to food access.

Food insecurity is defined by the United States Department of Agriculture (USDA) as limited or uncertain access to adequate food. It is a significant public health issue linked to malnutrition, chronic disease and functional limitations.(6) After a decade-long decline, U.S. food insecurity rose from 10.5% in 2021 to 13.5% in 2023, affecting 18.0 million households.(7) Arizona has one of the highest rates of food insecurity in the western United States. Nearly one in six individuals (14.4%) of the state’s population experienced food insecurity in 2023.(8) Long COVID may exacerbate this burden by compounding pre-existing vulnerabilities and intensifying socioeconomic inequities.(1,4)

There is a clear relationship between food insecurity and long COVID when examining the socioeconomic elements that contribute to both issues. While the role of food insecurity in immune function and recovery has been explored,(9) less attention has been paid to whether long COVID increases the risk of food insecurity. Understanding this relationship is key due to long COVID’s potential to cause reduced earning capacity, and prolonged care needs. This study uses data from the Arizona CoVHORT study to examine whether long COVID is associated with greater odds of food insecurity.

## METHODS

We used data from the Arizona CoVHORT, a prospective longitudinal cohort initiated in May 2020 to evaluate short- and long-term health impacts of SARS-CoV-2 infection among Arizona residents. Study procedures have been described previously.(10) Recruitment involved partnerships with state and local health departments during case investigations, outreach at testing and vaccination sites, and statewide postcard mailings. All residents were eligible regardless of infection status. Informed consent was obtained, and the University of Arizona Institutional Review Board approved the study (Protocol #2003521636).

Structured electronic surveys were administered via REDCap at baseline and quarterly thereafter. Data included sociodemographic, household income, education, SARS-CoV-2 testing, infection status, and food security questions. Participants with confirmed infection completed symptom surveys every six weeks until symptoms resolved, then resumed quarterly follow-up. Between January and May 2024, a supplemental survey was distributed to 4,794 eligible participants with prior COVID-19 infection, assessing food insecurity, barriers to care, and financial stressors. This analysis included 2,415 adults (≥18 years) meeting the following criteria: (1) confirmed positive SARS-CoV-2 test, (2) documented infection date, (3) at least one symptom survey ≥6 months post-infection, and (4) completion of the supplemental survey.

A matched cohort design was applied to reduce confounding. The analytic sample matched participants based on gender (male, female, nonbinary), age (±10 years), household income (≤$35,000; $35,001–$75,000; >$75,000), and timing of the first food insecurity survey (±6 months). Non-long COVID participants were prioritized for matching based on greater completeness of food insecurity assessments.

The primary exposure was long COVID status. The outcome was food insecurity, measured at each follow-up using a validated two-item screener adapted from the USDA Household Food Security Survey.(11) For this analysis, we included assessments between 15- and 33-months post-infection and classified participants as “ever food insecure” (≥1 report) or “never food insecure.” Descriptive statistics summarized characteristics by long COVID status, and group differences were assessed using chi-square tests. Associations between long COVID and food insecurity were estimated using conditional logistic regression, adjusting for gender, education, and income. Odds ratios (ORs) with 95% confidence intervals (CIs) were reported. Statistical significance was set at p < 0.05. Analyses were conducted using R version 4.4.1.

## RESULTS

Of the entire matched population, (n=2415), 22.4% (n=652) were identified as having long COVID. The long COVID population were predominantly white (92.3%), non-Hispanic, females with a median age of 51 years and average annual household income of $75,000 at baseline. The matching procedure achieved excellent balance on age, gender, income, and timing of the first food insecurity survey, with no statistically significant differences observed between long COVID positive and long COVID negative participants (all p > 0.05). Distributions of unmatched demographic characteristics, including race, ethnicity, and education, were also comparable across groups. Of the 2,415 matched participants, 224 (9.3%) were food insecure. Among the 652 participants with long COVID, 73 (12%) reported food insecurity, whereas among the 1,763 participants without long COVID, 151 (8.6%) reported food insecurity. To account for temporal variation, participants were matched on the timing of their first food insecurity survey (±6 months). Conditional logistic regression analysis accounted for the matched design and revealed a strong statistically significant association between long COVID and food insecurity (Table 1). The unadjusted model indicated 37% increased odds of food insecurity among individuals with long COVID (OR = 1.37; 95% CI: 1.00–1.87; p = 0.05), and this association showed 47% higher odds of food insecurity among participants with long COVID after further adjustment (OR = 1.47; 95% CI: 1.06–2.04; p = 0.02) (Table 1).

**Table 1.** Association Between long COVID and Food Insecurity Among Arizona CoVHORT Participants (n = 2,415): Arizona, May 2020-July 2024.

| Model | Predictor variable | Unadjusted OR (95% CI) | p-value | Adjusted OR (95% CI) | P-value |
| --- | --- | --- | --- | --- | --- |
| Conditional Logistic regression | Long COVID | 1.37 (1.00 - 1.87) | 0.05 | 1.47 (1.06 - 2.04) | 0.02* |

*Note: OR = odds ratio; CI = confidence interval. Adjusted models control gender, income, and education; \* $P < 0.05$ .*

## DISCUSSION

In this matched sub cohort of the Arizona CoVHORT, long COVID was significantly associated with increased odds of food insecurity. These findings are consistent with national and international evidence linking long COVID to socioeconomic hardship. A recent study utilizing data from the 2022 National Health Interview Survey (NHIS) reported a statistically significant association between long COVID and food insecurity, particularly among low-income U.S. adults.(12) Similarly, the PAMPA cohort in Brazil reported a significant association between long COVID and food insecurity(9), suggesting global relevance. In addition, findings show that individuals with long COVID were significantly more likely to be unemployed than those who had recovered from COVID-19 without long-term symptoms.(4) These disruptions in employment increases vulnerability to food insecurity among individuals with long COVID.

Strengths of this study include the matched design, which reduced confounding, the large, prospective cohort, and the use of a validated two-item screener, enhancing reliability and potential generalizability. Limitations include reliance on self-reported measures, which may introduce recall bias, and the potential for residual confounding from unmeasured variables such as household composition or social support.

## Data Availability

All data produced in the present work are contained in the manuscript.

## ACKNOWLEDGEMENTS

The authors wish to thank the participants of the Arizona CoVHORT study for their time and contributions. We also acknowledge the support of the research staff and colleagues who assisted with data collection, management, and interpretation.

