## Supplemental Table 1 for "Long COVID and the Increased Risk of Food Insecurity Among Participants in Arizona CoVHORT: A matched cohort study"

**Supplementary data**

| Supplementary Table 1: Demographics of matched long COVID positive and long COVID negative participants: Arizona, May 2020 – June, 2024 | | | | |
| --- | --- | --- | --- | --- |
|  | Long COVID + (n = 652) | | Long COVID - (n = 1763) | |
| Matched demographics | | | | |
| **Variables** | n | % | n | % |
| **Age (Median)** | 51.23 |  | 51.23 |  |
| **Gender** |  |  |  |  |
| Female | 489 | 75.0 | 1290 | 73.2 |
| Male | 154 | 23.6 | 461 | 26.1 |
| Non-binary | 5 | 0.8 | 8 | 0.5 |
| **Income** |  |  |  |  |
| Less than $10,000 | 5 | 0.8 | 15 | 0.9 |
| $10,000 to less than $20,000 | 10 | 1.5 | 36 | 2.0 |
| $20,000 to less than $25,000 | 12 | 1.8 | 29 | 1.6 |
| $25,000 to less than $35,000 | 22 | 3.4 | 56 | 3.2 |
| $35,000 to less than $50,000 | 54 | 8.3 | 180 | 10.2 |
| $50,000 to less than $75,000 | 127 | 19.5 | 287 | 16.3 |
| More than $75,000 | 422 | 64.7 | 1160 | 65.8 |
| Unmatched demographics | | | | |
| **Race** |  |  |  |  |
| White | 602 | 92.3 | 1615 | 91.6 |
| Black/African American | 7 | 1.1 | 19 | 1.1 |
| Mixed race | 16 | 2.5 | 48 | 2.7 |
| Asian | 12 | 1.8 | 39 | 2.2 |
| American Indian | 5 | 0.8 | 17 | 1.0 |
| **Ethnicity** |  |  |  |  |
| Non-Hispanic | 578 | 88.7 | 1581 | 89.7 |
| Hispanic or Latino | 69 | 10.6 | 169 | 9.6 |
| **Education** |  |  |  |  |
| High school | 10 | 1.5 | 23 | 1.3 |
| College (1-3 years) | 105 | 16.1 | 224 | 12.7 |
| College (4 + years) | 199 | 30.5 | 551 | 31.3 |
| Postgraduate | 337 | 51.7 | 964 | 54.7 |
