## Supplemental Table 2 for "Long COVID and the Increased Risk of Food Insecurity Among Participants in Arizona CoVHORT: A matched cohort study"

| Supplementary Table 2: Demographics of food secure and food insecure participants from the matched population (n = 2415): Arizona, May 2020 – June, 2024 | | | | |
| --- | --- | --- | --- | --- |
|  | Food Insecure (n = 224) | | Food Secure (n = 2191) | |
| **Variables** | n | % | n | % |
| **Age (Median)** | 51.23 |  | 51.23 |  |
| **Gender** |  |  |  |  |
| Female | 185 | 82.6 | 1728 | 78.9 |
| Male | 34 | 15.2 | 605 | 27.6 |
| Non-binary | 4 | 1.8 | 9 | 0.4 |
| **Income** |  |  |  |  |
| Less than $10,000 | 10 | 4.5 | 13 | 0.6 |
| $10,000 to less than $20,000 | 19 | 8.5 | 38 | 1.7 |
| $20,000 to less than $25,000 | 13 | 5.8 | 38 | 1.7 |
| $25,000 to less than $35,000 | 14 | 6.3 | 68 | 3.0 |
| $35,000 to less than $50,000 | 51 | 22.8 | 215 | 9.8 |
| $50,000 to less than $75,000 | 60 | 26.8 | 407 | 18.6 |
| More than $75,000 | 58 | 25.9 | 1570 | 71.7 |
| **Race** |  |  |  |  |
| White | 200 | 89.3 | 2159 | 98.6 |
| Black/African American | 7 | 3.1 | 22 | 1.0 |
| Mixed race | 7 | 3.1 | 62 | 2.8 |
| Asian | 6 | 2.7 | 50 | 2.3 |
| American Indian | 1 | 0.4 | 21 | 1.0 |
| **Ethnicity** |  |  |  |  |
| Non-Hispanic | 187 | 83.5 | 2104 | 96.1 |
| Hispanic or Latino | 34 | 15.2 | 228 | 10.4 |
| **Education** |  |  |  |  |
| High school | 7 | 3.1 | 31 | 1.4 |
| College (1-3 years) | 61 | 27.2 | 306 | 14.0 |
| College (4 + years) | 81 | 36.2 | 729 | 33.3 |
| Postgraduate | 75 | 33.5 | 1281 | 58.5 |
